# Phenotypic spectrum of *FAM47E*-*SHROOM3* haplotype composition in a general population sample

**DOI:** 10.1101/2024.03.22.24304731

**Authors:** Dariush Ghasemi-Semeskandeh, Eva König, Luisa Foco, Nikola Dordevic, Martin Gögele, Johannes Rainer, Markus Ralser, Dianne Acoba, Francisco S. Domingues, Dorien J. M. Peters, Peter P. Pramstaller, Cristian Pattaro

## Abstract

Genome-wide association studies identified a locus on chromosome 4q21.1, spanning the *FAM47E*, *STBD1*, *CCDC158*, and *SHROOM3* genes, as associated with kidney function markers. Functional studies implicated *SHROOM3*, encoding an actin-binding protein involved in cell shaping, into podocyte barrier damage. Despite the locus was also found associated with electrolytes, hematological and cardiovascular traits, systematic explorations of functional variants across all the genes in the locus are lacking.

We reconstructed haplotypes covering the whole locus on 12,834 participants to the Cooperative Health Research in South Tyrol (CHRIS) study, using genotypes imputed on a whole-exome sequencing reference panel of a subsample of 3,422 participants. Haplotypes included 146 exonic and intronic variants over the four genes and were tested for association with 73 serum, urine and anthropometric traits, 172 serum metabolite and 148 plasma protein concentrations using linear regression models.

We identified 11 haplotypes with 2% to 24% frequency. Compared to the most common haplotype, most haplotypes were associated with higher levels of the creatinine-based estimated glomerular filtration rate and lower serum magnesium levels. The second most common haplotype (12% frequency) was additionally associated with lower dodecanoyl-, hydroxyvaleryl- and tiglyl-carnitine serum concentrations. A haplotype of 4% frequency was also associated with lower red blood cell count, hemoglobin, and hematocrit levels. A haplotype of 2% frequency was associated with serum glutamine and putrescine concentrations. Cluster analysis revealed distinct groups of traits and of haplotypes.

The *FAM47E*-*SHROOM3* locus exhibits haplotype variability that corresponds to marked pleiotropic effects, implicating the existence of population subgroups with distinct biomarker profiles.

## Introduction

A genetic locus on chromosome 4q21.1 has been prominently associated with multiple markers of kidney function, including the estimated glomerular filtration rate based on serum creatinine (eGFRcrea) or cystatin C^1–6^ and albuminuria.^7^ The genetic associations lie within two recombination hotspots that embed a high linkage disequilibrium (LD) region spanning four genes: Family With Sequence Similarity 47 Member E (*FAM47E*); Starch Binding Domain 1 (*STBD1*); Coiled-Coil Domain Containing 158 (*CCDC158*); and the first exons of the Shroom Family Member 3 (*SHROOM3*).

Follow-up functional studies have mainly focused on *SHROOM3*, which encodes a PDZ-domain-containing protein regulating cell shape, neural tube formation^8^ and epithelial morphogenesis. SHROOM3 is involved in maintaining normal podocyte structure,^9^ particularly through binding of the TCF7L2 transcription factor.^10^ *Shroom3* modulates the actomyosin network which maintains podocyte architecture.^11^ Variants altering the *SHROOM3* actin-binding domain may cause podocyte effacement and glomerular filtration barrier impairment.^9^ Furthermore, *Shroom3* knockdown causes albuminuria and podocyte foot process effacement in mice as well as defective lamellipodia formation in podocytes and disrupted slit diaphragms in rat glomerular epithelial cells.^12^ *SHROOM3* alterations may also implicate craniofacial alterations^13,14^ and cardiac defects.^15^ Less attention has been dedicated to the other genes in the locus.

Beyond kidney function, genetic variants at this locus are associated with diverse phenotypes, including platelets,^16^ hemoglobin (HGB),^16^ serum magnesium,^17^ and Parkinson’s disease.^18^ The alleles associated with lower eGFRcrea are also associated with lower urine albumin-to-creatinine ratio (UACR),^7^ despite glomerular *Shroom3* knockdown should induce albuminuria.^19^

Altogether, this evidence suggests that, while *SHROOM3* is a promising molecular target for chronic kidney disease (CKD) prevention,^10^ the genetic variability of the whole *FAM47E-SHROOM3* locus warrants more extensive investigations to comprehensively characterize the corresponding phenotypic spectrum. This may also clarify whether the previously reported, apparent mediatory role of serum magnesium on the association between eGFRcrea and the locus^20^ reflects real biological mechanisms or rather the joint genetic regulation of the two traits.

To investigate the presence of distinct genetic profiles involving diverse phenotypic manifestations, we conducted a haplotype analysis of genetic variants in the *FAM47E-SHROOM3* region imputed from a whole-exome sequencing (WES) panel, in an Alpine population-based study. Notoriously, haplotype analysis does not illuminate causal mechanisms but instead, it is a population genetic tool to explore a locus’s heterogeneity in population. Reconstructed haplotypes were tested for association against 73 clinical traits, 172 target serum metabolites, and 148 plasma protein markers.

## Methods

### Study sample

We analyzed data from the Cooperative Health Research in South Tyrol (CHRIS) study, conducted in South Tyrol, Italy, between 2011 and 2018.^21^ Following overnight fasting, 13,393 participants underwent early morning blood drawing, urine collection, anthropometric measurements, and blood pressure measurements.^22^ Health and lifestyle information was gathered through computer-based standardized questionnaire-based interviews. The analysis overview is given through a flowchart in **Figure 1A**.

**Figure 1.**
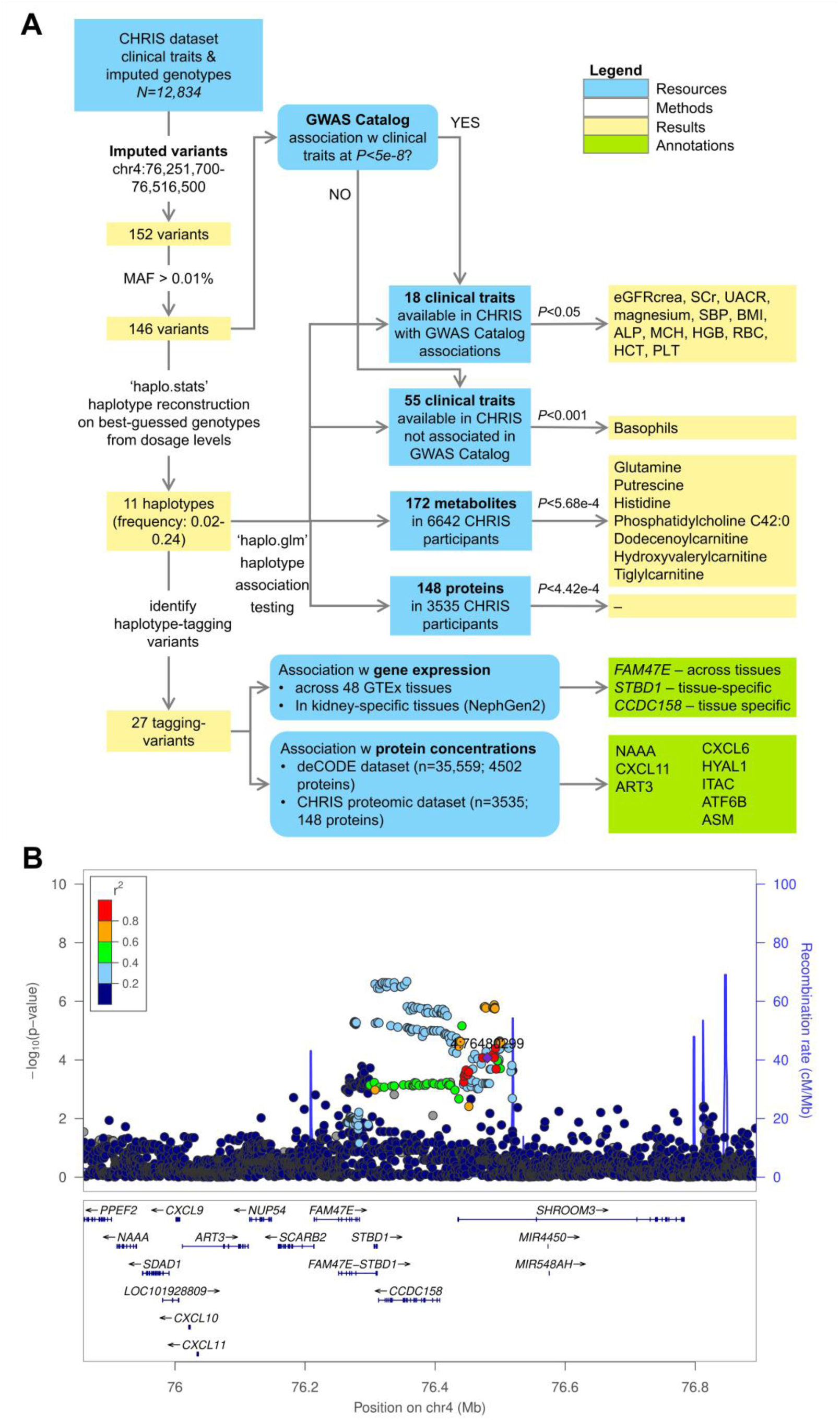
Analysis setting. **Panel A.** Analysis flowchart. **Panel B.** Regional association plot depicting associations between variants in and around the *FAM47E*-*SHROOM3* locus in the CHRIS study, reflecting a similar association pattern as that identified by a recent CKDGen GWAS meta-analysis^4^: the most associated SNP in the CKDGen analysis is highlighted in purple. SNP positions are referred to the NCBI Build 38. Plot generated with LocusZoom v1.4^57^.

### Genomics

All DNA samples were genotyped on the Illumina HumanOmniExpressExome or Omni2.5Exome arrays and called with GenomeStudio v2010.3 with default settings on GRCh37, lifted to GRCh38 via CrossMap v0.6.5. Variants with GenTrain score <0.6, cluster separation score <0.4, or call rate <80% were considered technical failures and discarded. Variants present on both arrays were submitted to further quality control (QC) and removed if monomorphic or not in Hardy-Weinberg equilibrium (*P*<10*^-^*^6^). Samples with <0.98 call rate were removed.

Samples from a subset of 3840 participants underwent WES (xGen® Exome Research Panel v1.0; McDonnell Genome Institute, Washington University). Data processing, read alignment and QC were conducted as detailed previously.^23,24^ We retained 3422 samples with a post-QC mean target coverage of 68.4X.

WES data were used as a population-specific reference panel for genotype imputation onto the whole CHRIS sample as reported previously.^23^ Based on 181 WES samples excluded from the reference panel for testing purposes, we observed excellent imputation quality between imputed genotypes and sequenced hard-calls (median Pearson’s correlation=0.99).^23^

### Clinical, metabolomics, and proteomics traits

Information on genotype data and clinical traits was available for 12,834 participants. Clinical traits included blood and urinary markers, diastolic and systolic blood pressure (SBP), and body mass index (BMI; **Suppl. Tab. 1**). eGFRcrea was estimated with the race-free CKD-EPI equation using the R package ‘nephro’ v1.3.0.^25^ Laboratory assay effects^26^ were addressed through quantile normalization as detailed previously.^20^ Missing values were imputed to the median.

Targeted metabolomics analysis involved a subset of 7252 participants, whose serum samples were analyzed with the AbsoluteIDQ p180 kit (Biocrates Life Sciences AG, Innsbruck, Austria). Normalization and QC of the 188 measured metabolites are described elsewhere.^27^ To increase sample homogeneity, pregnant women and individuals of non-European descent were excluded. Metabolites with >20% missing data were excluded. Missing values were imputed to the median, otherwise. QC left 172 high-quality metabolites available on 6642 samples (**Suppl. Tab. 1**).

On a subset of 4087 participants we measured 148 plasma proteins, using mass-spectrometry-based scanning SWATH.^28^ Data generation and processing was described elsewhere.^29^ After merging with the genetic data, 3535 participants remained for the analysis (**Suppl. Tab. 1**).

### Haplotype association analysis

The region of interest on chromosome 4 was bounded by two recombination peaks at positions 76,251,700 and 76,516,500, encompassing 152 WES-based imputed variants spanning *FAM47E*, *FAM47E*–*STBD1*, *CCDC158* and *SHROOM3* **(Figure 1B)**. Retaining 146 variants with minor allele frequency>0.0001 and with imputation quality index Rsq>0.3 (median Rsq=0.87; **Suppl. Tab. 2**), haplotype reconstruction and regression analysis was conducted using the *haplo.glm* function of the R package ‘haplo.stats’ v1.8.9, exploiting an expectation-maximization algorithm for haplotype inference^30^ (**Supplementary Methods**). Alleles were aligned on the major allele as the reference. Haplotypes with <0.02 frequency were collapsed into a rare-haplotype category. We fitted linear association models on the inverse normal transformation of each trait, metabolite, and protein, including haplotypes as predictors and adjusting for age, sex, and the first 10 genetic principal components (PCs), estimated on the genotyped autosomal variants, to control for population structure.

Among the 73 considered clinical traits, 18 traits have been previously reported with genome-wide significant associations with variants in the locus (GWAS Catalog interrogation at https://www.ebi.ac.uk/gwas/ on 23-Aug-2023; **Figure 2A**). These traits were tested for association with the haplotypes at the significance level α=0.05, considering the strong prior evidence of association. For the remaining clinical traits, the 172 metabolites, and the 148 proteins, we set α at 0.05/50=0.001, 0.05/88=5.68×10^-4^ and 0.05/113=4.42×10^-4^, respectively, where 50, 88, and 113 were the number of independent PCs necessary to explain 95% of each dataset’s variability (analyses conducted with the *prcomp* function in the R package ‘stats’ v4.3.0).

**Figure 2.**
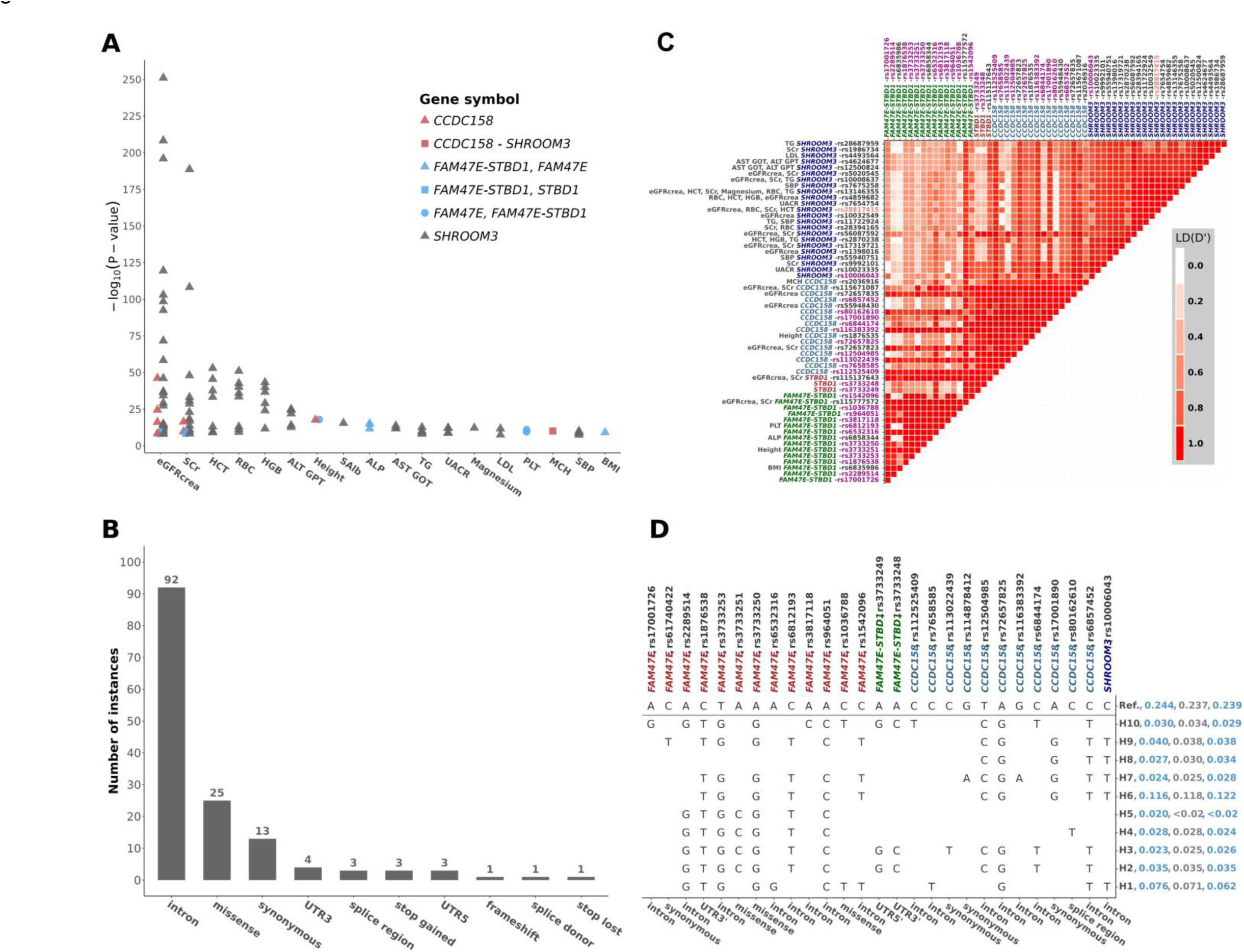
Characteristics of the variants included in the *FAM47E*-*SHROOM3* region on chromosome 4 (76,251,700-76,516,500 bp). **Panel A.** Minus log_10_ P-values of the significant associations between any of the 146 variants used for haplotype reconstruction and 18 GWAS Catalog traits that are also present in the CHRIS study. Colors and shapes of the dots are used to distinguish the different genes the variants belong to. Traits: creatinine-based estimated glomerular filtration rate, eGFRcrea; serum creatinine, SCr; hematocrit, HCT; red blood cell count, RBC; hemoglobin, HGB; alanine transaminase, ALT GPT; height; serum albumin, SAlb; alkaline phosphatase, ALP; aspartate aminotransferase, AST GOT; triglycerides, TG; urine albumin-creatinine ratio, UACR; serum magnesium; low-density lipoprotein (LDL) cholesterol; platelet count, PLT; mean corpuscular hemoglobin, MCH; systolic blood pressure, SBP; and body mass index, BMI. **Panel B.** Barplot of the most severe consequences of the 146 variants identified for haplotype reconstruction. **Panel C**. Linkage disequilibrium pattern of the *FAM47E*-*SHROOM3* locus, based on the D’ statistic. Included are variants associated with complex traits from previous genome-wide association studies and haplotype-tagging variants. The labels on x- and y-axis are annotated by the corresponding gene symbol and RSID. On the vertical axis, we also report the associated complex trait, as per GWAS Catalog interrogation. Variants’ RSID color coding indicates: the most associated variant, rs28817415, with eGFRcrea ^5^ (orange); 25 of the 27 haplotype-tagging variants (two variants were not represented in the LD reference panel) (purple); and 31 variants for which there was a GWAS Catalog genome-wide significant association with a trait among those included in the CHRIS study (black). **Panel D**. Distribution of the 11 reconstructed haplotypes, identified by 27 tagging variants with their functional consequences given on the x-axis. Haplotype frequencies are reported right of the haplotype label for the three analyzed subsamples in the order: all individuals with clinical traits; those with also metabolites measurements; those with additional protein measurements.

### Cluster analysis

We performed hierarchical clustering of the z-scores obtained from the significant associations between haplotypes and traits and metabolites. Similarity was based on the Euclidean distance and clustering implemented based on the ‘Ward D2’ approach via *hclust* in the ‘stats’ R package. We applied the Silhouette method to the non-scaled z-scores to determine the optimal number of clusters for haplotypes and traits, using the *fviz_nbclust and hcut* functions in the R package ‘factoextra’ v1.0.7, allowing a maximum of 8 and 18 clusters, respectively.

### Variant annotation

Genetic variants were annotated with the Ensembl Variant Effect Predictor v100.2 (http://www.ensembl.org/info/docs/tools/vep/index.html), predicting the most severe consequences with the ‘split-vep’ plugin. LD between WES-imputed variants selected for haplotype analysis and previously reported common variants associated with the traits of interest was assessed through the D′ statistic, which reflects the underlying haplotype diversity,^31^ estimated using PLINK v1.9.^32^

We queried the haplotype-tagging variants in the European ancestry datasets of the GTEx Consortium v8 database (https://gtexportal.org/home/; 10-Aug-2023) across 48 tissues (n=65 to 573 samples per tissue) and in NephQTL2^33^ (6-Feb-2024), to test association with the expression of the four genes in the locus (*P*<5×10^-8^), and across 4502 whole blood protein GWAS summary results available in ^34^ to identify protein quantitative trait loci (pQTLs) at *P*<5×10^-8^ (n=35,559; https://decode.com/summarydata; 11-Oct-2023).

## Results

### Characterization of the FAM47E-SHROOM3 genomic variants

The 12,834 participants (54.3% females) had a median age of 46 years and median eGFRcrea level of 92.9 ml/min/1.73m^2^; 3.6% had eGFRcrea<60 ml/min/1.73m^2^ and 5.9% had UACR>30 mg/g (**Table 1**). The sample appeared as an extract of the general population, with no particularly prevalent clinical aspects (**Suppl. Tab. 1**).

**Table 1.** Main characteristics of the study sample. Data are described as median (interquartile range) or number of cases (percentage), as appropriate. Additional characteristics are described in Supplementary Table 1.

| Participants' characteristics | Group of traits and sample size |  |  |
| --- | --- | --- | --- |
|  | Clinical traits<br>(n=12,834) | Metabolites<br>(n=6,642) | Proteins<br>(n=3,535) |
| Age, years | 46 (31 – 57) | 46 (32 – 58) | 46 (32 – 58) |
| Females | 6,969 (54.3%) | 3,650 (55.0%) | 1,977 (55.9%) |
| eGFR<sub>crea</sub>,<br>ml/min/1.73 m<sup>2</sup> | 92.9 (81.7 – 104.1) | 92.2 (81.1 – 103.5) | 92.2 (80.8 – 103.3) |
| eGFR<sub>crea</sub> < 60<br>ml/min/1.73 m<sup>2</sup> | 460 (3.6%) | 269 (4.1%) | 143 (4.1%) |
| UACR > 30 mg/g | 752 (5.9%) | 393 (5.9%) | 225 (6.4%) |
| HbA1c > 6.5% | 189 (1.5%) | 97 (1.5%) | 54 (1.5%) |
| Hypertension* | 2,023 (15.8%) | 1,002 (15.1%) | 536 (15.2%) |

\* systolic blood pressure >140 mmHg or diastolic blood pressure >90 mmHg

Within the locus we identified 146 WES-imputed variants. They were largely intronic, but also included missense variants as well as synonymous, stop-gain, splicing, and other types of functional variants (**Figure 2B, Suppl. Tab. 2**). The variants were all in strong-to-perfect LD with common variants previously associated with common traits (**Figure 2C**). GWAS catalog interrogation showed the marked locus’ pleiotropic nature (**Figure 2A**; **Suppl. Fig. 1**).

Haplotype reconstruction identified 11 haplotypes (H1 to H11) with ≥2% frequency (**Suppl. Fig. 2**), which were uniquely tagged by 27 of the 146 variants (**Figure 2D**): rs3733251, rs3733250, and rs1036788 are missense variants in *FAM47E* and rs80162610 is a splice variant at *CCDC158*. Similar haplotype distributions were observed in the metabolomics and proteomics subsamples (**Figure 2D**).

The 27 haplotype-tagging variants were not associated with *SHROOM3* expression (**Figure 3A**; **Suppl. Tab. 3**). rs2289514, rs1876538, rs3733253, rs964051, and the missense variant rs3733250 at *FAM47E* were associated with *FAM47E* expression across most tissues. The remaining variants were associated with the expression of at least one among *FAM47E*, *CCDC158*, and *STBD1* in different tissues. No variant was a kidney-specific eQTL for the four investigated genes in the GTEx. NephQTL2 database interrogation did not identify any genome-wide significant eQTLs in the glomerular and tubulointerstitial tissues (**Suppl. Tab. 4**).

**Figure 3.**
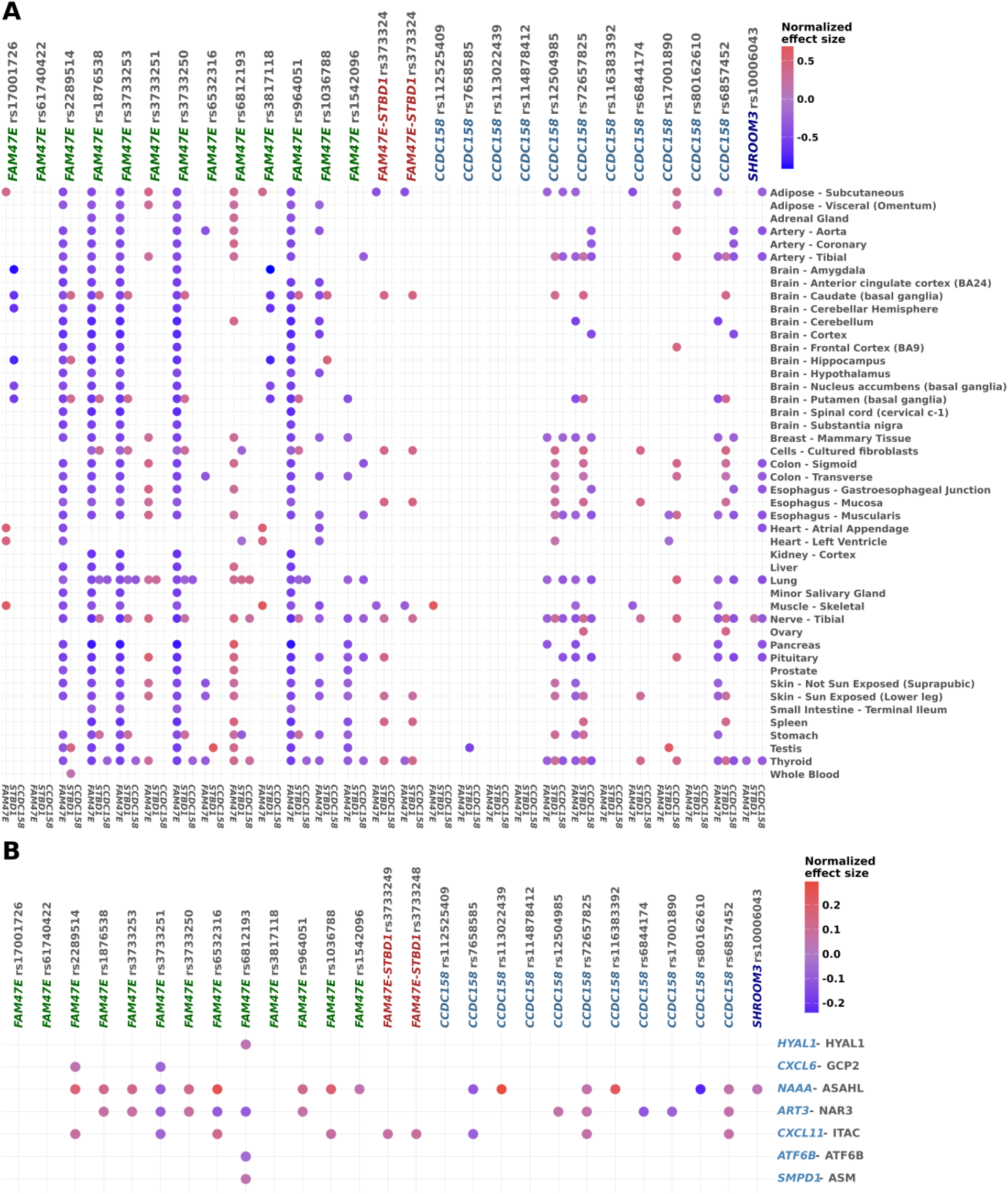
Association of the 27 haplotype-tagging variants with gene expression and protein levels. **Panel A**: Normalized effect size of association between the 27 haplotype-tagging variants and expression of *FAM47E*, *STBD1*, and *CCDC158* (horizontal axis; genes grouped by variant) across 46 tissues (vertical axis) retrieved from the GTEx v8 dataset. Uterus and vagina tissues and *SHROOM3* were omitted for the lack of significant results. **Panel B**: Normalized effect size for association between the 27 haplotype-tagging variants (horizontal axis) and protein concentrations (vertical axis) retrieved from the deCODE dataset ^34^. Listed are only proteins with a significant association with at least one variant.

The proteins encoded by the four genes in the locus were not included in the CHRIS or in the deCODE plasma proteomic datasets [36]. However, in the latter, we observed associations with proteins (**Figure 3B**) whose encoding genes are also located on chromosome 4q21.1, immediately to the left of the recombination peak next to *FAM47E*: the ADP-ribosyltransferase 3 (NAR3) encoded by *ART3*, the N-acylethanolamine acid amidase (NAAA) encoded by the homonymous gene, and the C-X-C motif chemokine ligand 11 (CXCL11; see **Figure 1B** for localization). Twenty-two of the 27 haplotype-tagging variants were associated with at least one of those three proteins. Specifically, *FAM47E* missense variant rs3733251 was associated with all three proteins and with the C-X-C motif chemokine ligand 6 (CXCL6), whose encoding gene seats on the contiguous 4q13 cytoband. Other variants associated with all three adjacent proteins were rs6532316 in *FAM47E* and rs72657825 and rs6857452 in *CCDC158.* NAAA was associated with most variants, including the *SHROOM3* intronic variant rs10006043. Variant rs6812193 in *FAM47E* was additionally associated with sphingomyelin phosphodiesterase 1 (SMPD1), hyaluronidase 1 (HYAL1), and the activating transcription factor 6 beta (ATF6B), whose encoding genes are in different chromosomes.

### Haplotype association analyses and hierarchical clustering

Haplotypes were first tested for association with the 18 traits for which there was a previously reported association with single variants at the locus (**Table 2, Suppl. Tab. 5**). Compared to the reference haplotype, haplotype H1 was associated with higher SBP and lower levels of mean corpuscular hemoglobin (MCH), HGB and magnesium. H4 was associated with lower serum magnesium and creatinine levels as well as higher eGFRcrea, UACR, and BMI levels. H5 was associated with lower alkaline phosphatase (ALP). H6, the second most common haplotype (frequency=11.6%), was associated with lower serum creatinine (*P*=4.92×10^-6^) and platelet count (PLT) and higher eGFRcrea (*P*=0.004). H7 was associated with lower serum magnesium and creatinine (*P*=0.044). H8 was associated with lower serum creatinine (*P*=0.001) as well as higher eGFRcrea (*P*=2.72×10^-4^), UACR (*P*=0.003) and SBP (*P*=0.019). H9 was associated with lower HGB, red blood cell count (RBC), hematocrit, magnesium, serum creatinine and higher eGFRcrea. H10 was associated with lower magnesium levels. For the 55 remaining traits without prior evidence of association with variants in the locus, we identified an additional association between H4 and lower basophil levels (*P*=4.18×10^-4^), after multiple testing control.

**Table 2.** Statistically significant* haplotype associations with blood, urine, anthropometric, and metabolic traits in the CHRIS study. No significant association was observed with proteins.

| Group | Haplotype | Trait | Effect (SE)** | P-value |
| --- | --- | --- | --- | --- |
| Clinical traits | H1 | SBP | 0.054 (0.023) | 0.017156 |
|  |  | MCH | -0.063 (0.027) | 0.018654 |
|  |  | HGB | -0.044 (0.021) | 0.037743 |
|  |  | Magnesium | -0.056 (0.027) | 0.041443 |
|  | H4 | Basophils | -0.145 (0.041) | 4.18E-04 |
|  |  | Magnesium | -0.139 (0.041) | 6.74E-04 |
|  |  | eGFRcrea | 0.089 (0.029) | 0.002221 |
|  |  | BMI | 0.118 (0.039) | 0.002364 |
|  |  | Serum creatinine | -0.094 (0.032) | 0.003637 |
|  |  | UACR | 0.085 (0.039) | 0.027938 |
|  | H5 | ALP | -0.101 (0.049) | 0.039383 |
|  | H6 | Serum creatinine | -0.084 (0.018) | 4.92E-06 |
|  |  | eGFRcrea | 0.048 (0.017) | 0.003950 |
|  |  | PLT | -0.052 (0.023) | 0.021181 |
|  | H7 | Magnesium | -0.106 (0.043) | 0.014985 |
|  |  | Serum creatinine | -0.069 (0.034) | 0.043883 |
|  | H8 | eGFRcrea | 0.113 (0.031) | 2.72E-04 |
|  |  | Serum creatinine | -0.111 (0.034) | 0.001136 |
|  |  | UACR | 0.122 (0.041) | 0.003278 |
|  |  | SBP | 0.084 (0.036) | 0.019310 |
|  | H9 | HGB | -0.069 (0.027) | 0.010716 |
|  |  | RBC | -0.069 (0.030) | 0.022546 |
|  |  | HCT | -0.063 (0.028) | 0.025566 |
|  |  | eGFRcrea | 0.056 (0.025) | 0.026760 |
|  |  | Magnesium | -0.075 (0.035) | 0.032351 |
|  |  | Serum creatinine | -0.054 (0.028) | 0.049913 |
|  | H10 | Magnesium | -0.125 (0.040) | 0.001810 |
| Metabolites | H1 | Phosphatidylcholine diacyl C42:0 | 0.134 (0.038) | 4.39E-04 |
|  | H3 | Histidine | -0.215 (0.061) | 4.28E-04 |
|  | H6 | Dodecenoylcarnitine | -0.116 (0.032) | 2.82E-04 |
|  |  | Hydroxyvalerylcarnitine *** | -0.122 (0.032) | 1.31E-04 |
|  |  | Tiglylcarnitine | -0.113 (0.032) | 4.48E-04 |
|  | H10 | Glutamine | -0.196 (0.051) | 1.08E-04 |
|  |  | Putrescine | -0.195 (0.053) | 2.26E-04 |

\*Statistical significance was set at $\alpha = 0.05$ for traits with prior evidence of association and $\alpha = 0.05$ to the number of principal components explaining 95% of the set variability for traits and metabolites with no prior evidence of association (see Methods). \*\*Effects are expressed in terms of standard deviations as all traits were normalized using the inverse-normal transformation (see Methods) \*\*\*Also known as Methylmalonylcarnitine

Haplotypes were also associated with serum metabolites (**Table 2**; **Suppl. Tab. 6**): H1 with higher phosphatidylcholine C42:0 (*P*=4.39×10^-4^); H3 with lower histidine (*P*=4.28×10^-4^); H10 with lower glutamine (*P*=1.08×10^-4^) and putrescine (*P*=2.26×10^-4^). H6 was associated with lower dodecenoylcarnitine (*P*=2.82×10^-4^), hydroxyvalerylcarnitine (*P*=1.31×10^-4^), and tiglylcarnitine (*P*=4.48×10^-4^) concentrations. **Suppl. Fig. 3** outlines all associations between haplotypes, clinical traits, and metabolites. After multiple testing control, no significant association between haplotypes and the 148 targeted plasma proteins was identified (**Suppl. Tab. 7**).

For haplotypes associated with both clinical traits and metabolites, we repeated the haplotype-metabolite association tests adjusting for the clinical traits (**Table 3**): haplotype effects on metabolites remained generally unchanged, indicating independent effects, except for H6 effect on carnitines that was attenuated by serum creatinine adjustment, potentially indicating common regulatory mechanisms.

**Table 3.** Covariate-adjusted haplotype-metabolite association models. Association models between each haplotype and the associated metabolite (Table 2) were adjusted for the traits associated with the same haplotype to assess potential mediation.

| Haplotype | Phenotype | Covariate | Effect (SE) | P-value |
| --- | --- | --- | --- | --- |
| **H1** | Phosphatidylcholine diacyl C42:0 | SBP | 0.135 (0.038) | 0.000340 |
|  |  | HGB | 0.134 (0.038) | 0.000437 |
|  |  | Magnesium | 0.133 (0.038) | 0.000458 |
| **H6** | Dodecenoylcarnitine | Serum creatinine | -0.097 (0.031) | 0.001950 |
|  |  | eGFRcrea | -0.106 (0.032) | 0.000792 |
|  |  | PLT | -0.117 (0.032) | 0.000264 |
|  | Hydroxyvalerylcarnitine<br>(Methylmalonylcarnitine) | Serum creatinine | -0.108 (0.032) | 0.000645 |
|  |  | eGFRcrea | -0.114 (0.032) | 0.000322 |
|  |  | PLT | -0.122 (0.032) | 0.000129 |
|  | Tiglylcarnitine | Serum creatinine | -0.097 (0.032) | 0.002170 |
|  |  | eGFRcrea | -0.103 (0.032) | 0.001130 |
|  |  | PLT | -0.115 (0.032) | 0.000336 |
| **H10** | Glutamine | Magnesium | -0.197 (0.051) | 0.000104 |
|  | Putrescine | Magnesium | -0.195 (0.053) | 0.000239 |

### Cluster analyses of significant trait-haplotype associations

Hierarchical clustering of the haplotype effects on the significantly associated 13 clinical traits and 7 metabolites, identified 6 clusters of traits, according to the Silhouette method (**Figure 4**; **Suppl. Fig. 4**). Cluster 1 included red blood cells (HGB, hematocrit, RBC, MCH) and immune traits (basophils). Cluster 2 grouped together putrescine, glutamine, and histidine. Cluster 3 included serum creatinine and magnesium. Cluster 4 involved a combination of clinical traits (PLT, BMI) and metabolites (dodecenoylcarnitine). Cluster 5 included carnitines hydroxyvalerylcarnitine and tiglylcarnitine. Cluster 6 included traits associated with kidney function (eGFRcrea, UACR), blood pressure (SBP), liver transaminases (ALP) and phosphatidylcholine C42:0. When clustering by haplotype, the Silhouette method identified two clusters, one grouping H1, H3, H6, H9 and H10 and a second one including H2, H4, H7, and H8. Altogether, the identified cluster structure suggests the existence of distinct genetic and correspondingly phenotypic characteristics at *FAM47E*-*SHROOM3*, where individuals with specific haplotypes present specific phenotypic and metabolic signatures.

**Figure 4.**
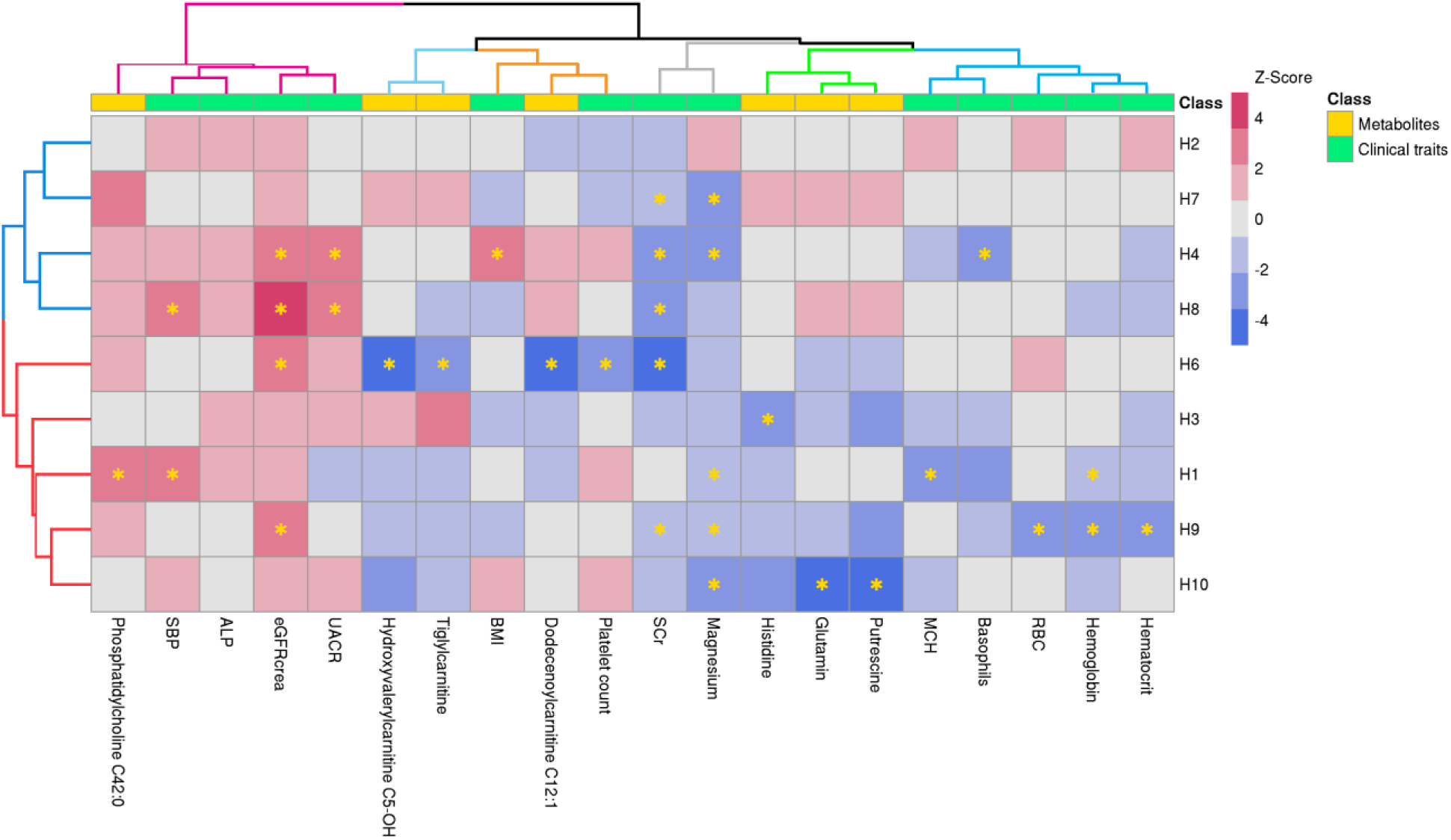
Hierarchical cluster analysis of z-scores from the associations between haplotypes with any of the 13 clinical traits and 7 metabolites that were associated with at least one haplotype (indicated with *), in the subset of study participants with metabolites measurements available. Haplotype H5 was excluded as it did not reach the 2% frequency in this subsample.

## Discussion

Our comprehensive investigation of 11 haplotypes derived from 146 WES-imputed variants and evaluated across 393 clinical traits, metabolites, and proteins, highlights haplotypic profiles at *FAM47E*-*SHROOM3* associated with distinct phenotype manifestations. We extend previous studies on the role of *SHROOM3* on kidney function regulation,^10^ by additionally showing that haplotypes spanning *SHROOM3* include missense variants at *FAM47E*, which seems the most relevant gene in the haplotype after *SHROOM3*, raising the possibility of complex interactions. Given its function of promoting histone methylation, *FAM47E* might epigenetically regulate *SHROOM3* transcription. *STBD1* is involved in glycogen metabolism and protein is absent in the kidney, while *CCDC158* is strictly testis specific.

The diffuse association of most haplotypes with both lower serum magnesium and higher eGFRcrea suggests that the previously observed mediation between the two traits^20^ more likely reflects their joint regulation by genes in the locus. Association of haplotypes H4 and H8 with lower serum creatinine and higher UACR and eGFRcrea at the same time, is compatible with reduced muscle mass causing lower circulating creatinine and thus higher eGFRcrea and lower urinary creatinine excretion increasing UACR. Nevertheless, H8 association with higher SBP is consistent with the increased risk of incident hypertension in albuminuric individuals.^35^ Similar results were observed for H4, despite non-significant association with higher SBP. Given H4 and H8 do not share any alleles at the 27 tagging variants, their similar characteristics might reflect LD with functional variants outside the reconstructed haplotypes.

Haplotype H6 was associated with lower serum creatinine and lower acylcarnitines dodecenoylcarnitine, hydroxyvalerylcarnitine, and tiglylcarnitine concentrations. Acylcarnitines transport fatty acids from the cytosol into the mitochondria to produce energy through beta-oxidation.^36^ They are freely filtered by the kidney and excreted in the urine.^37^ As kidney function decreases, serum acylcarnitines should increase.^38^ This was observed in CKD patients exhibiting high serum dodecenoylcarnitine, hydroxyvalerylcarnitine and tiglylcarnitine concentrations linked to low eGFR.^37^ This aligns with our findings, suggesting that H6 might confer nephroprotection through carnitine reduction. Alternatively, H6 might be in LD with *SHROOM3* alleles conferring sustained structural integrity of the podocytes, resulting in better filtration capacity lowering both creatinine levels and free acylcarnitines concentrations.

Haplotypes H9 and H1 were associated with lower levels of red blood cell traits. The kidney produces erythropoietin, which is key to red blood cell production.^39^ Lower kidney function could lead to less or defective red blood cells but connection with H9 is difficult as the haplotype is associated at the same time with higher eGFRcrea.

H3 carriers had lower histidine concentrations and statistically non-significant lower putrescine and glutamine concentrations. Similarly, H10 carriers showed significantly lower putrescine and glutamine and non-significant lower histidine. Histidine is an anti-inflammatory and antioxidant factor. In CKD patients, low histidine concentrations are associated with energy wasting, inflammation, oxidative stress, and mortality,^40^ and histidine supplementation is beneficial.^41^ Kidney filtration plays a role in putrescine concentrations:^42^ dialyzed patients have lower serum putrescine than non-dialyzed patients.^43^ A study on diabetic mice has postulated that putrescine from ornithine catabolism activates mTOR signaling,^44^ leading to podocyte loss by regulating autophagy, oxidative stress, and endoplasmic reticulum stress.^45^ Under glycolytic blockage, podocytes use amino acids as oxidative phosphorylation substrates to produce energy.^44^ Less putrescine in H10 carriers might reflect well-functioning podocytes and glomeruli following proper *SHROOM3* function. However, such protective effect contradicts the simultaneous negative association between H3 and histidine and between H10 and glutamine. Glutamine, the most abundant amino acid in humans, synthesized by most tissues and involved in numerous metabolic pathways,^46^ is a fundamental precursor of glutathione. Under fasting or starvation, glutamine serves gluconeogenesis, helping the liver to maintain blood glucose levels following glycogen-store shortages.^47^ In rats, glutamine supplementation protects against STZ-induced renal injury and prevents downregulation of the kidney injury molecule-1 (KIM-1), neutrophil gelatinase-associated lipocalin (NGAL), TGF-β1, and collagen-1 mRNA expressions.^48^ In diabetic patients, glutamine supplementation decreases glycemia through increased glucagon-like peptide 1 (GLP-1) secretion.^49^ By presenting lower glutamine concentrations, H10 carriers might have more difficulty in rebalancing glucose metabolism in the event of glucose deficiency. The absence of associations of H3 and H10 with clinical traits limits further interpretation.

We modeled haplotypes within two recombination hotspots at positions 76,251,700 and 76,516,500 on chromosome 4q21.1 that clearly delineate the genetic locus associated with eGFR in all GWAS reported so far. Prokop et al.^10^ showed that variants within those two peaks are in LD with variants outside the peaks such as *SHROOM3* P1244L, located downstream the second peak and associated with high CKD risk in Eastern Asia.^10^ They also observed that the locus is associated with *TCF7L2*, a chromosome 10 transcriptional factor with a broad phenotypic spectrum.^50^ Our annotation analyses extend these observations, showing that several haplotype-tagging variants are also associated with proteins (NAR3, CXCL11, and NAAA) whose encoding genes are located immediately upstream the recombination hotspot adjacent to *FAM47E*, between chromosome 4 positions 75,910,655 and 76,114,048. These associations might reflect underlying longer haplotypes spanning those genes as well. NAR3 is encoded by *ART3*, which is specifically expressed in the mesangium of the glomerulus.^51^ It warrants further investigation whether joint involvement of *SHROOM3* on podocytes and *ART3* on the mesangium is possible and if that might implicate relevant kidney phenotypes. *CXCL11* is a proinflammatory chemokine implicated in kidney disease induced by interferon signaling.^52^ Urine CXCL11 correlates with diabetic kidney disease progression^53^ and is upregulated in the glomeruli of nephrotic syndrome patients carrying *APOL1* high risk variants.^54^ In mice with acute glomerular inflammation, genetic deletion of *CXCL11* receptor *Cxcr3* attenuates glomerulosclerosis and albuminuria.^55^ NAAA is a proinflammatory protein emerging as a promising target in mouse models of parkinsonism,^56^ without evident links to kidney function. Other associated proteins, encoded by genes in other chromosomes (*SMPD1*, *HYAL1*, *ATF6B*, and *CXCL6*), may reflect either *FAM47E* transcriptional activity or biological consequences of the proteins encoded by the genes tagged by the haplotypes.

The main strength of our analysis was the availability of WES data from a subsample which we used to impute exonic variants in the whole study sample of >12,000 individuals, enabling the reconstruction of haplotypes with frequencies as low as 2% in the population across clinical traits, metabolites, and proteins. Limitations are also present. The proteomics panel included mostly highly abundant plasma proteins, none of which resulted associated with haplotypes at this locus. The successful single-variant query of external proteomic datasets suggests that haplotype associations with those same proteins might be identified should individual-level data become available. Despite the large sample size, haplotypes are multicategory variables that easily generate data sparseness, eroding statistical power. Finally, consistent with GWAS studies that identified significant associations with complex traits at this locus, we focused our investigation on the region bounded by the two recombination hotspots at chromosome 4 positions 76,251,700 and 76,516,500: given most *SHROOM3* exons fall outside this segment, this constraint has probably limited the possibility to contextualize *SHROOM3* with the other genes. On the other hand, extrapolating haplotypes at arbitrary distance outside the borders would have increased data sparseness.

In conclusion, our investigation revealed the presence of distinct genetic profiles at *FAM47E*-*SHROOM3* associated with heterogeneous phenotypic and metabolic combinations that warrant dedicated investigation.

## Ethics approval and consent to participate

The Ethics Committee of the Healthcare System of the Autonomous Province of Bolzano-South Tyrol approved the CHRIS baseline protocol on 19 April 2011 (21-2011). The study conforms to the Declaration of Helsinki, and with national and institutional legal and ethical requirements. All participants included in the analysis gave written informed consent.

## Acknowledgements

The CHRIS study is conducted in collaboration between the Eurac Research Institute for Biomedicine and the Healthcare System of the Autonomous Province of Bolzano-South Tyrol. Investigators thank all study participants, the general practitioners of the Val Venosta/ Vinschgau district, and the staff of the Silandro/Schlanders hospital and of the Autonomous Province of Bolzano-South Tyrol Healthcare System for their support and collaboration. They also thank the study team in Silandro/Schlanders, the CHRIS Biobank personnel, and all Institute for Biomedicine colleagues who contributed to the study. Extensive acknowledgement is reported at: https://www.eurac.edu/en/institutes-centers/institute-for-biomedicine/pages/acknowledgements. CHRIS Bioresource Research Impact Factor (BRIF) code: BRIF6107. The authors thank the Department of Innovation, Research and University of the Autonomous Province of Bolzano-South Tyrol for covering the Open Access publication costs.

## Author contributions

Conceptualization of the research project: C.P., D.G.

Recruitment and study management: M.G., P.P.P., C.P.

Bioinformatics: D.G., E.K., N.D.

Data quality control and harmonization: M.G., J.R., M.R., L.F.

Statistical Analysis: D.G.

Results interpretation: D.G., C.P., M.G., L.F., D.A.

Manuscript drafting: D.G., C.P.

Manuscript critical revision: All authors

## Data availability

The data used in the current study can be requested with an application to at the Eurac Research Institute for Biomedicine.

## Supplementary Methods

Haplotypes were reconstructed using the expectation-maximization algorithm implemented in the R package ‘haplo.stats’ v1.8.9 with the following parameter setting: n.try=2 (number of times to try to maximize the log-likelihood); insert.batch.size=2 (number of loci to be inserted in a single batch); max.haps.limit=4e6 (maximum number of haplotypes for the input genotypes); and min.posterior=1e-5 (minimum posterior probability for a haplotype pair, given the input genotypes).

---

**Supplementary Figure 1.**
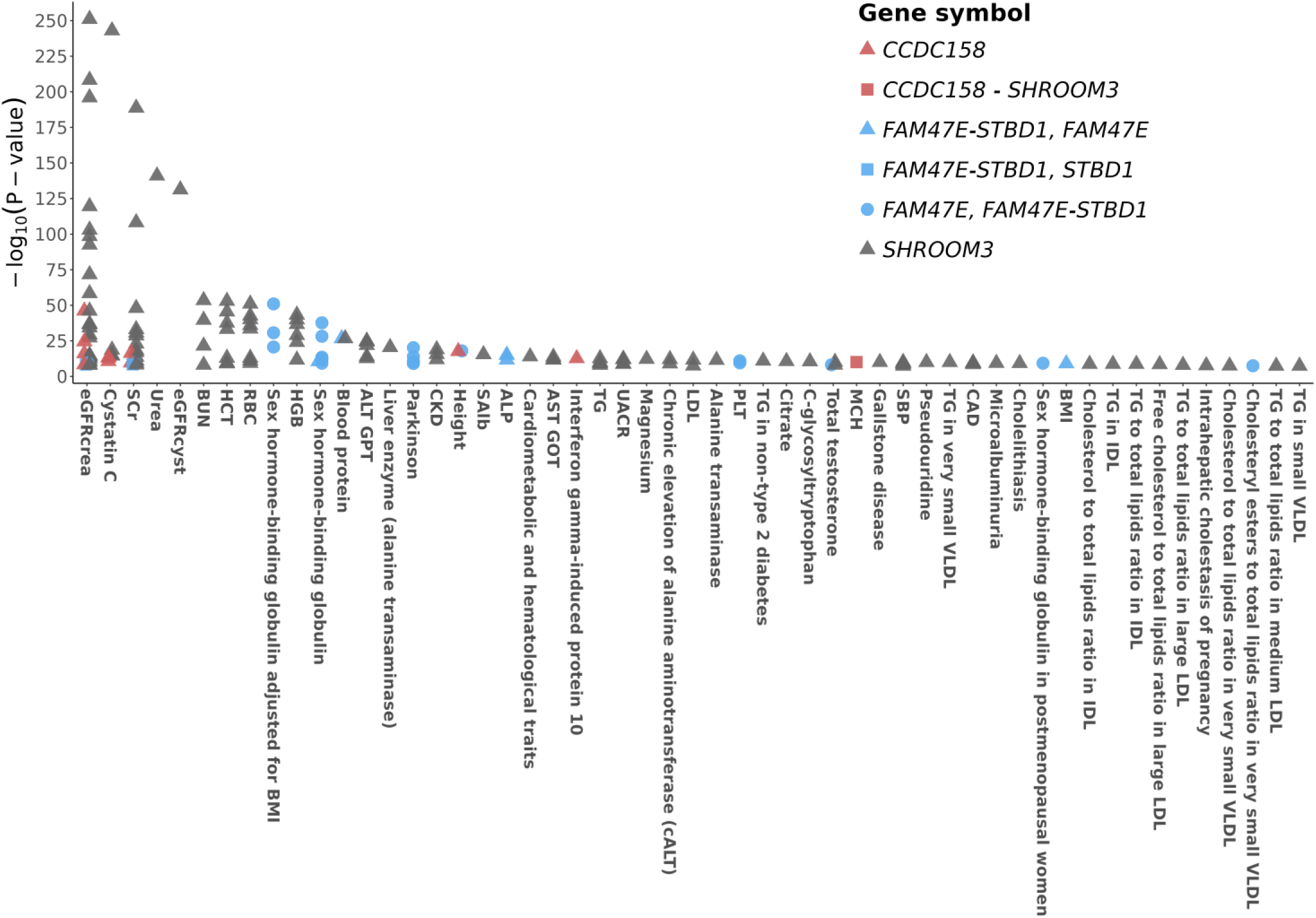
Interrogation of the 146 variants in association with any complex trait in the GWAS Catalog. Minus log_10_ P-values of the significant associations are presented. Only genome-wide significant associations (P-value<5×10^-8^) are presented. Colors and shapes of the dots are used to distinguish the different genes to which the variants belong.

**Supplementary Figure 2.**
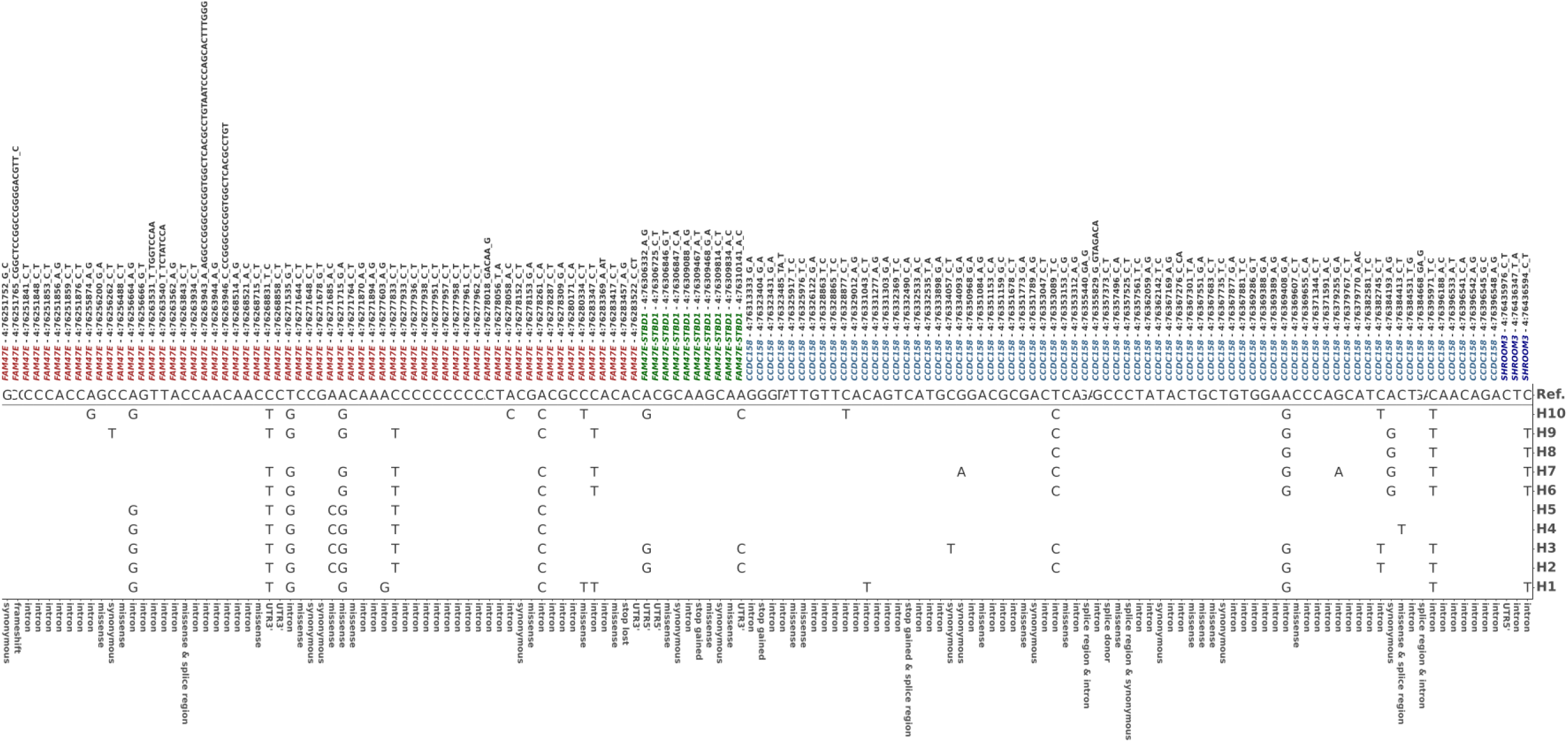
Haplotypes reconstructed based on the 146 available WES-imputed variants. On top, variants are listed by gene name – chromosomal position – alleles. At the bottom, variants’ functional consequences. Middle panel: the most common, reference haplotype is provided in its entirety (first line). Other haplotypes are presented by listing only the alleles that are different from the reference haplotype. When the allele is the same as in the reference haplotype, the letter is not displayed.

**Supplemental Figure 3.**
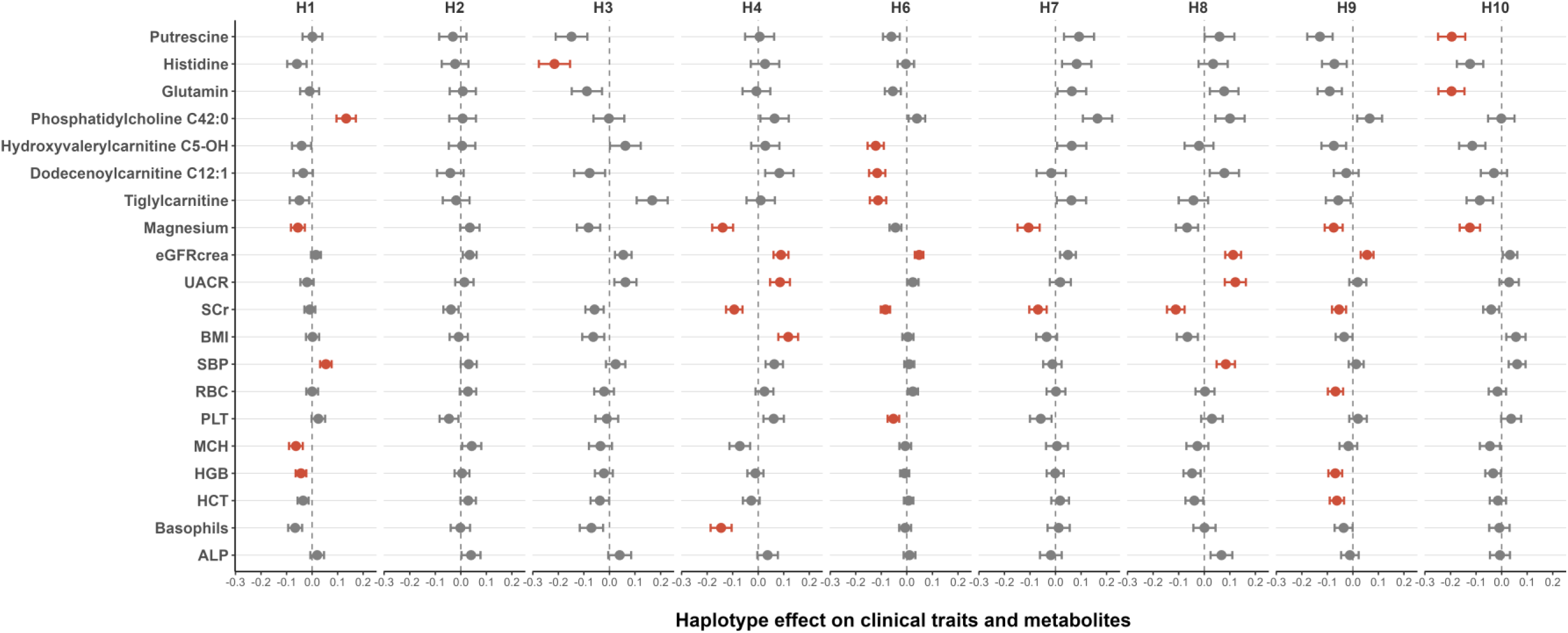
Haplotype association analysis results. Displayed are the effect coefficients and their 95% confidence intervals from the associations between haplotypes with the 13 clinical traits and 7 metabolites that were associated with at least one haplotype, in the subset of participants with metabolites measurements available. Haplotype 5 (H5) was excluded as it did not reach the 2% frequency in this subsample.

**Supplemental Figure 4.**
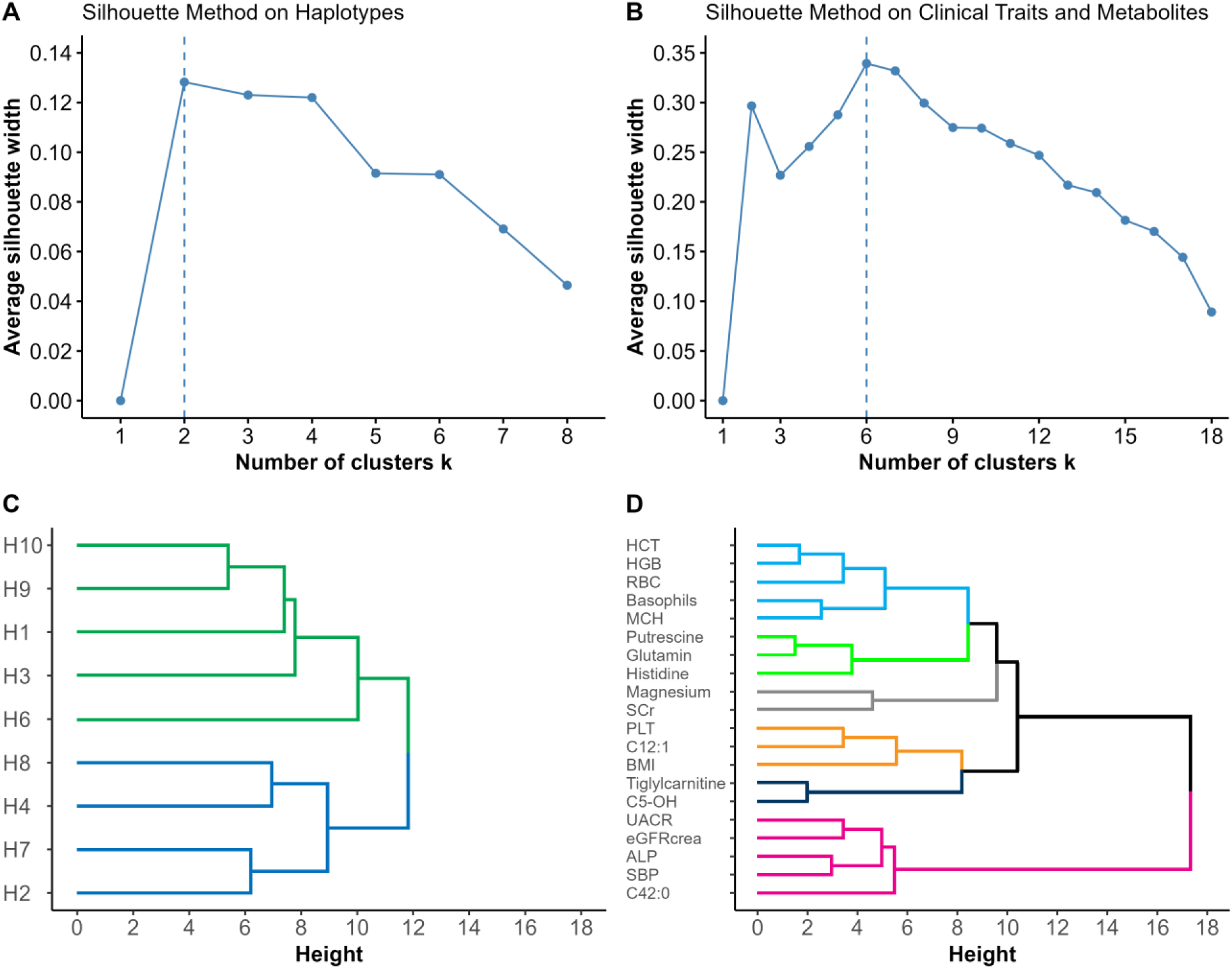
Cluster analysis of the standardized effects of haplotypes on significant clinical traits and metabolites. **Panel A**. Results of the Silhouette method applied to the clustering of haplotypes: the number of clusters (x-axis) is plotted against the average silhouette width (y-axis). **Panel B**. Results of the Silhouette method applied to the clustering of traits: the number of clusters (x-axis) is plotted against the average silhouette width (y-axis). **Panel C**. Hierarchical clustering of haplotypes (listed and clustered on the y-axis). **Panel D**. Hierarchical clustering of clinical traits and metabolites (listed and clustered on the y-axis). Colors are used to identify the clusters.

